# Load and muscle dependent changes in triceps surae motor unit firing properties and motor unit firing-torque relationships in individuals with non-insertional Achilles tendinopathy

**DOI:** 10.1101/2024.08.27.24312381

**Authors:** Ignacio Contreras-Hernandez, Deborah Falla, Michail Arvanitidis, Francesco Negro, David Jimenez-Grande, Eduardo Martinez-Valdes

## Abstract

Non-insertional Achilles tendinopathy (NIAT) induces morpho-mechanical changes to the Achilles tendon (AT). However, evidence on how triceps surae motor unit firing properties are influenced by altered tendon mechanics in NIAT is limited. This study investigated motor unit firing properties (mean discharge rate (DR), recruitment and de-recruitment thresholds, and discharge rate variability (COVisi)), motor unit firing-torque relationships (cross-correlation coefficient between cumulative spike train (CST) and torque, and neuromechanical delay), and neural drive distribution (connectivity strength and functional networks) of the medial gastrocnemius (MG), lateral gastrocnemius (LG), and soleus (SO) muscles during isometric plantarflexion contractions at 10%, 40%, and 70% maximal voluntary contraction (MVC) using high-density surface electromyography on 26 individuals with NIAT and 25 healthy controls. Furthermore, AT’s morpho-mechanical properties (thickness, cross-sectional area, length and stiffness) were assessed via ultrasound imaging. NIAT individuals showed reduced tendon stiffness and increased thickness (p<0.01). Motor unit properties changed in a load and muscle-dependent manner. LG DR increased (p=0.002) and de-recruitment threshold decreased (p=0.039) at 70%MVC in the NIAT group compared to controls. The CST-torque cross-correlation coefficient of the LG decreased at 10%MVC (p<0.0001) and increased at 70%MVC (p=0.013) in the NIAT group. Connectivity strength for the 0-5 Hz and 5-15 Hz frequency bands decreased (p<0.01) in the NIAT group at 10%MVC. This study shows that individuals with NIAT exhibit load-dependent changes in motor unit firing properties, motor unit-torque relationships, and neural drive distribution to the triceps surae. These alterations may be due to muscle-specific compensations for the modified mechanical properties of the AT.

**KEY POINTS:**

- Individuals with non-insertional Achilles tendinopathy (NIAT) have changes of the neural drive to the lateral gastrocnemius (LG) muscle and altered contribution of the LG to the net plantarflexion torque.
- Individuals with NIAT show a more uneven distribution of neural drive to the triceps surae muscle at low force levels, characterized by reduced intermuscular coherence between the medial and lateral gastrocnemius in the 0-5 Hz and 5-15 Hz bands compared to the control group.
- Our findings support the idea that the LG may have a central role in the pathophysiology of this condition, possibly affecting the load transmission to the Achilles tendon (AT).

## INTRODUCTION

Non-insertional Achilles tendinopathy (NIAT) is a common debilitating tendon disorder in the lower extremity, characterized by an insidious onset of pain accompanied by swelling, stiffness and thickening in the midportion of the Achilles tendon (AT) (Maffulli *et al*., 1998; van Dijk *et al*., 2011; Bahari *et al*., 2023; Sara *et al*., 2023). NIAT can affect athletes and the general population, causing disability, functional impairment and decreasing performance (Kvist, 1994; de Jonge *et al*., 2011). It’s estimated that more than half (55-65%) of the AT problems occur about 2-7 cm proximal to superior border of the calcaneus (Järvinen *et al*., 2005). The etiology of the NIAT remains debated (Maffulli *et al*., 2020); however, excessive loading of the tendon is considered the major causative factor (Leadbetter, 1992). Recent evidence suggests that uneven activation of the triceps surae muscle may influence the magnitude and distribution of AT load, stress and strain (O’Neill *et al*., 2015; Debenham *et al*., 2017); therefore, deficits in the performance of this muscle group has been identified as a key factor for the development of NIAT (O’Neill *et al*., 2019). In humans, plantarflexion is primarily accomplished by the triceps surae muscle group, which comprises the medial gastrocnemius (MG), lateral gastrocnemius (LG) and soleus (SO) muscles (Héroux *et al*., 2014). The resultant force generated by these muscles is then transmitted to the AT, which links the triceps surae muscle to the calcaneus (Crouzier *et al*., 2020). The AT is composed of three subtendons, arising from each of the triceps surae muscles (Crouzier *et al*., 2020); together, these subtendons exhibit a twisted structure, with the deep layer of the tendon formed by fibers from the LG and SO, and the superficial layer formed by fibers from the MG and SO, with a wide range of rotation of the subtendons between individuals (Szaro *et al*., 2009). Ultrasound imaging during isometric contractions has revealed nonuniform displacements within the Achilles tendon, consistent with an uneven distribution of load across the tendon (Arndt *et al*., 1998; Clark & Franz, 2018). Additionally, 3D models suggest that the nonuniform distribution of load within the AT is largely due to the distribution of force among the MG, LG, and SO muscles (Handsfield *et al*., 2017). Consequently, it has been proposed that an imbalance of the force produced by these three muscles may cause an heterogenous load within the tendon, contributing to the development of NIAT(Bojsen-Møller & Magnusson, 2015; Hug & Tucker, 2017).

The morpho-mechanical properties of the Achilles tendon in individuals with NIAT have been studied extensively. The morphological changes include an increase in tendon thickness, cross-sectional area (CSA), and volume (Shalabi *et al*., 2004; Arya & Kulig, 2010; Child *et al*., 2010; Grigg *et al*., 2012; Docking & Cook, 2016) and the mechanical changes comprise an increase in longitudinal and transverse strain, hysteresis, and lower stiffness and Young’s modulus compared to healthy tendons under the same tensile load (Arya & Kulig, 2010; Child *et al*., 2010; Wang *et al*., 2012; Chang & Kulig, 2015). Alterations in the AT morpho-mechanical properties are relevant because of their potential to modify the stress-strain patterns experienced by the AT during muscle contraction (Hansen *et al*., 2017), and muscle’s capacity to function within the force-length curve’s optimal region (Arya & Kulig, 2010), possibly influencing the triceps surae motor unit firing properties.

For the generation of movement, the central nervous system (CNS) must effectively coordinate the many degrees of freedom of the musculoskeletal system, considering the nonlinear features of muscles and the dynamic interactions between the segments of the body, as well as the interactions between the body and the environment (d’Avella & Bizzi, 2005). This complex task might be simplified by organizing a modular and hierarchical control (Full & Koditschek, 1999; Loeb *et al*., 1999; Bizzi *et al*., 2002). It has been hypothesized that the CNS uses a set of muscle synergies, the coherent activation in space or time of a group of muscles, as output modules. Thus, supraspinal and afferent signals combine with a determined number of muscle synergies to generate a variety of muscle activation patterns (d’Avella & Bizzi, 2005). Based on this, it has been theorized that alterations in how the CNS control the coordination of the triceps surae muscles could create uneven loading of the AT and contribute to tendinopathy (Cook & Purdam, 2009). Although these muscles act as agonists that share the same common distal tendon, they exhibit anatomical, neurophysiological, and functional differences, indicating diverse functional roles (Héroux *et al*., 2014). These functional roles have been linked to unique properties in motor unit firing properties between muscles when performing different tasks (Héroux *et al*., 2014).

The force exerted by a muscle during a voluntary contraction depends, partly, on the recruitment and discharge rate of motor units (Enoka & Duchateau, 2017). Hence, alterations in motor performance may be the consequence of the inability of the CNS to increase the recruitment and/or the discharge rate of the pool of motor units (Enoka & Duchateau, 2017). Within this framework, the most reliable approach to investigate how the CNS modulates the recruitment and discharge firing rates of the triceps surae motor units during a voluntary contraction is through the individual analysis of the MG, LG, and SO muscles (Hug *et al*., 2021; Fernandes *et al*., 2023). To the best of our knowledge, only one study has investigated the motor unit firing rates properties of the triceps surae muscles in individuals with NIAT. Fernandes et al. 2021 reported no differences in the discharge rate of the LG between a group of runners with NIAT and a control group at low forces; however, they observed that in the NIAT group the DR of the LG remained unchanged as the target torque increased from 10% to 20% MVC, suggesting that the neural drive to the LG is affected in individuals with NIAT (Fernandes *et al*., 2023). However, the authors only assessed motor unit firing parameters during low-force isometric plantarflexion contractions in a limited group of participants, without examining variations in AT’s morpho-mechanical properties and the distribution of the neural drive received by the triceps surae. Considering the stress-strain properties of tendons, it is known that the rate of tendon deformation changes according to the load placed on the tendon (Kongsgaard *et al*., 2011). Furthermore, the pain experienced by individuals with NIAT increases with load (Magnusson *et al*., 2010). Therefore, studying these properties across a wider range of torques is essential to understand how different loading conditions affect tendon behavior and motor unit firing patterns across the triceps surae muscle. Moreover, since common modulations in firing rate reflect the effective neural drive received by muscles (i.e., common synaptic input (Farina *et al*., 2014), it is crucial to examine these modulations between the different heads of the triceps surae and their relationship with the resultant torque. This information could provide insights into whether NIAT-induced changes in tendon structure might alter the contribution of each triceps surae muscle head to the produced torque.

We aimed to investigate triceps surae motor unit firing properties (mean discharge rate (DR), recruitment/de-recruitment thresholds and discharge rate variability (coefficient of variation of the interspike interval (COVisi)), motor unit firing-torque relationships (cross-correlation coefficient between cumulative spike train (CST) and torque, and neuromechanical delay (NMD)), and common motor unit firing modulations between triceps surae muscle heads (connectivity strength and muscular functional networks via intermuscular coherence) in individuals with NIAT at low, intermediate and high force levels during isometric plantarflexion contractions. In addition, morpho-mechanical properties of the AT (length, thickness, cross-sectional area (CSA) and stiffness) were determined using B-mode ultrasonography and shear-wave elastography (SWE). Our primary hypothesis is that NIAT-induced alterations of the AT’s morpho-mechanical properties would induce load and muscle-dependent changes in motor unit firing properties, motor unit firing-torque relationships and unbalanced distribution of the neural drive received by the triceps surae.

## METHODS

### Ethical approval

This study was approved by the Science, Technology, Engineering and Mathematics Ethical Review Committee, University of Birmingham, UK (ERN_20-0604A).

### Participants

Twenty-five healthy (17 males, 8 females, 28.60 ± 3.92 years, 74.00 ± 11.57 kg, 171.10 ± 9.22 cm) and twenty-six participants with NIAT (14 males, 12 females, 29.04 ± 8.46 years, 75.09 ± 17.20 kg, 173 ± 9.16 cm) were recruited from the University of Birmingham staff/student population and the local community via leaflets, e-mail, and social media.

Inclusion criteria included men or women aged 18 to 55 years old. This range was chosen to prevent the influence of ageing-related alterations in tendon properties, since previous studies have found that the AT has lower stiffness and Young’s modulus in older individuals (Lindemann *et al*., 2020). For the NIAT group inclusion criteria were as follows: presence of NIAT determined by an experienced physiotherapist based on defined clinical findings (VISA-A (Robinson *et al*., 2001), NRS (Numeral Rating Scale) (Alghadir *et al*., 2018) and pain for more than 3 months (Beyer *et al*., 2015)), physical examination and ultrasound imaging. Ultrasound imaging included assessing echoic pattern (focal hypoechoic and hyperechoic areas within the tendon) and tendon thickness (focal or diffuse thickening) (Bleakney & White, 2005). VISA-A scores less than 90 were considered as a reference to identify individuals with NIAT (Iversen *et al*., 2012). Due to the high variability in the NRS scores in individuals with NIAT(Rompe *et al*., 2009; Beyer *et al*., 2015), we considered individuals with an NRS score ≥ 2. Inclusion criteria for the healthy group included confirmation of a healthy tendon determined by the same physiotherapist through physical examination and ultrasound imaging.

The exclusion criteria for both groups were as follows: (1) current or history of chronic neurological, respiratory, or cardiovascular diseases, (2) systemic or inflammatory conditions including rheumatic, neuromuscular disorders, and malignancy, and (3) history of lower limb surgery. Specific exclusion criteria for the participants with NIAT were if they had undergone a corticosteroid injection in the Achilles tendon within the last 12 months or if they had been diagnosed with insertional Achilles tendinopathy. Specific exclusion criteria for the healthy group were history of NIAT, lower limb surgery or pain/injury in the lower limbs within the previous 6 months.

### Study design

This cross-sectional study was conducted from October 2021 to April 2023 at a laboratory within the Centre of Precision Rehabilitation for Spinal Pain (CPR Spine), University of Birmingham, UK. The study was conducted according to the declaration of Helsinki and participants signed written informed consent prior to participation. The guideline for Strengthening the Reporting of Observational Studies in Epidemiology (STROBE) was used to facilitate reporting (Vandenbroucke *et al*., 2007).

Participants made a single visit to the laboratory for the experimental session (2.5 hours). They were instructed to avoid any vigorous physical activity within 24 hours prior to testing. The assessed leg was randomized in the control group, and the leg with tendinopathy symptoms was evaluated in the NIAT group. In the case of individuals with bilateral symptoms, the most painful leg was assessed.

### Experimental setup and tasks

Experimental sessions included completing a battery of questionnaires, physical examination, ultrasonography of the AT, high-density surface electromyography (HD-sEMG) of the triceps surae muscles and torque recordings during plantarflexion isometric contractions. A representation of the experimental setup is shown in **Figure 1**.

**Figure 1.**
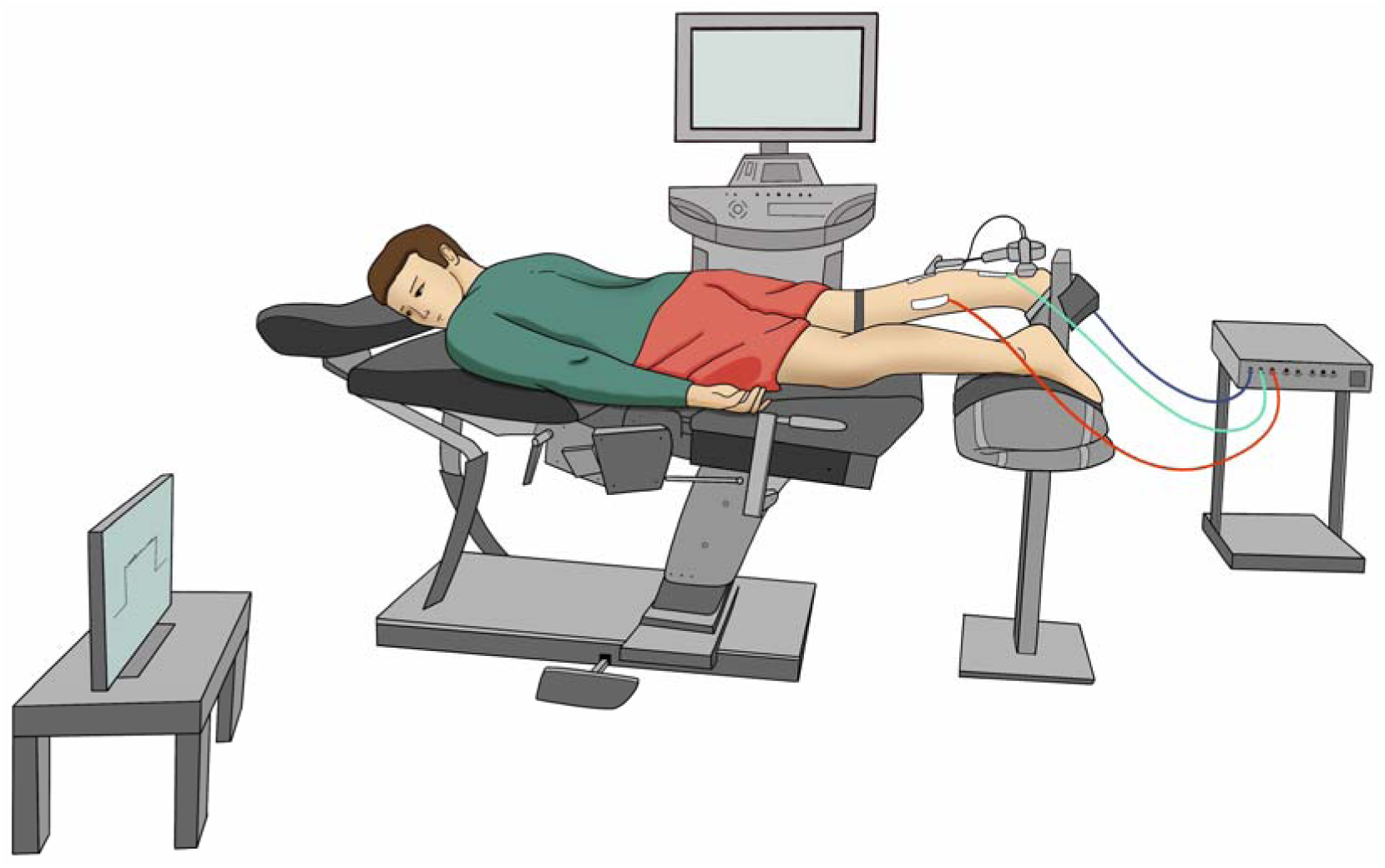
Representation of the experimental setup.

Anthropometric data (age, gender, weight, height, and foot dominance) was obtained across the participants. Foot preference in specific daily activities (foot dominance) was determined using a behavioral foot-preference inventory (Chapman *et al*., 1987). For the NIAT group, we obtained information regarding their symptoms (intensity of pain (NRS), duration of symptoms, and presence of bilateral symptoms). Then, all participants were asked to complete a battery of questionnaires, which included the Victorian Institute of Sports Assessment – Achilles Questionnaire (VISA-A) (Robinson *et al*., 2001), the International Physical Activity Questionnaire short form (IPAQ-SF) (Craig *et al*., 2003), Foot and Ankle Ability Measure (FAAM) (Martin *et al*., 2005), Pain Catastrophizing Scale (PCS) (Sullivan *et al*., 1995) and Tampa Scale for Kinesiophobia (TSK) (Kori, 1990). Then, participants lay prone on the chair of a Biodex System 3 dynamometer (Biodex Medical System), with their knees extended and their assessed foot tightly strapped on the footplate. The ankle was positioned in 0° of plantarflexion with the axis of the dynamometer aligned with the inferior tip of the lateral malleolus. The pelvis was stabilized with a strap to minimize compensatory movements. Ultrasonography was used to determine the length, thickness, and cross-sectional area of the AT (see procedure below). Then, the skin was shaved, cleaned, and prepared, and the electrodes grids were placed on the MG, LG, and SO muscles (see details below). Following the placement of the electrode grids, we assessed AT stiffness through SWE during rest conditions. HD-sEMG was used to confirm that the muscles were not active during these measurements as this could influence the estimation of stiffness.

Next, participants performed a warm-up protocol consisting of 3 submaximal isometric plantarflexion contractions at their perceived 30% of maximal voluntary force for 5 seconds with 30 seconds rest between contractions. The maximal voluntary contraction (MVC) was then determined during 3 isometric plantarflexion contractions of 5 seconds with 2 minutes rest between contractions (Martinez-Valdes *et al*., 2018) at 0° of plantarflexion. The highest MVC value was used as the reference maximal torque. Then, we measured the electromyographic activity of the MG, LG, and SO muscles during two isometric plantarflexion contractions at 10, 40 and 70% MVC (10% MVC/s ramp-up, 10 s hold, 10% MVC/s ramp-down and 30 s rest) with HD-sEMG. Visual feedback of the torque produced by the participant was provided through a computer monitor positioned 1 m in front of the participant. The order of the contractions at different target torque levels (10, 40, and 70% MVC) was randomized using a randomization app (Randomizer, https://play.google.com/store/apps/details?id=com.giannis.randomizer&hl=en_US). A schematic representation of the experimental session is shown in **Figure 2**.

**Figure 2.**
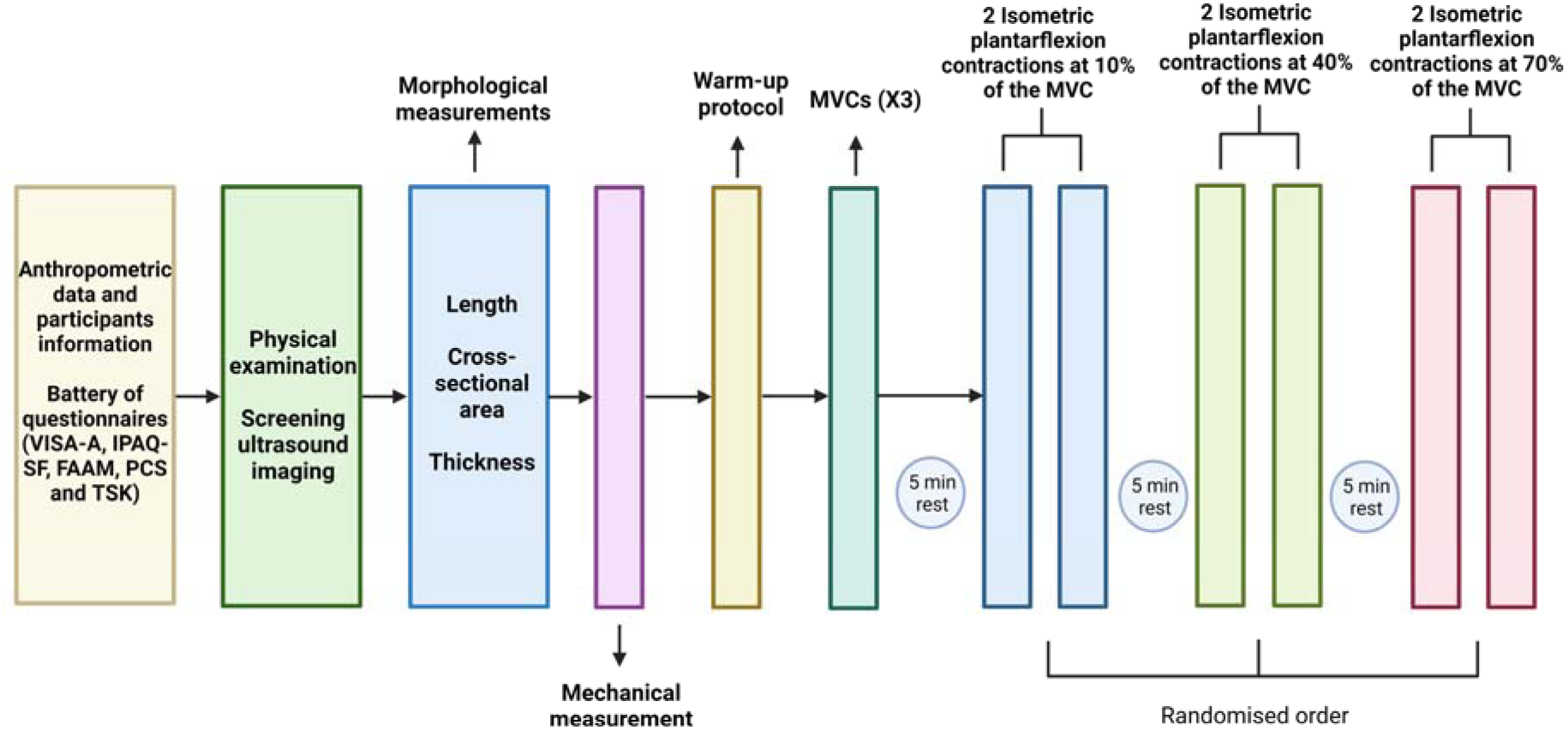
Schematic of the experimental procedure. The order of the contractions performed at each target torque (10, 40, and 70% MVC) was randomized. VISA-A, Victorian Institute of Sports Assessment-Achilles Questionnaire; IPAQ-SF, International Physical Activity Questionnaire short form; FAAM, Foot and Ankle Ability Measure; PCS, Pain Catastrophizing Scale; TSK, Tampa Scale for Kinesiophobia; MVC, maximum voluntary contraction.

### Ultrasonography

All ultrasound images were obtained using an ultrasound imaging device equipped with SWE (LOGIQ S8 GE Healthcare, Milwaukee, USA). Tendon thickness, length, and cross-sectional area were recorded in B-mode with a 16-linear array probe (50 mm, 4-15 MHz). SWE was recorded in elastography mode with a 9-linear array probe (44 mm, 2-8 MHz).

The morphological properties of the AT were determined using an adaptation of the protocol developed by Arya and Kulig (Arya & Kulig, 2010). The ultrasound probe was placed longitudinally over the posterior part of the heel with the ankle at 0° of plantarflexion, and the calcaneal notch was identified. Then, a fine wire (3.2 x 40 mm) was used under the probe to create an artifact in the ultrasound image. The artifact was aligned with the calcaneal notch and the corresponding point was marked on the skin with a marker (this mark represented the AT’s insertion). The ultrasound probe was moved proximally to identify the musculotendinous junction of the MG, and again, the wire was aligned with the musculotendinous junction of the MG and the corresponding point was marked on the skin. The distance between these two marks (calcaneal notch and musculotendinous junction of the MG represented the length of the AT). Subsequently, marks were made at 2, 4 and 6 cm above the AT’s insertion and these marks were used as reference to place the middle part of the probe in the longitudinal plane to obtain the thickness of the AT at 2, 4 and 6 cm of its insertion. Similarly, these reference marks were used to place the probe in the transversal plane and determine the cross-sectional area at 2, 4 and 6 cm of the AT’s insertion. Three ultrasound images were obtained in the longitudinal and transverse planes for each mark.

For the HD-sEMG electrode grid placement, a tape and marker were used to draw a line following the direction of the AT. This line and the distal musculotendinous junction of the MG were used as references for the positioning of the electrode grids. Briefly, for the MG electrode grid, a mark was made 10 cm above the distal musculotendinous junction of the MG and 4 cm medial to the line that followed the direction of the AT. Similarly, for the LG electrode grid, the leg was marked 10 cm above the distal musculotendinous junction of the MG and 4 cm lateral to the line that followed the direction of the AT. Likewise, for the SO electrode grid, the leg was marked 5 cm below the distal musculotendinous junction of the MG and 4 cm lateral to the line that followed the direction of the AT. The central electrodes of the HD-sEMG grids (electrode in row 7 and column 3) were placed on top of all these marks.

For the mechanical parameters (i.e., estimated stiffness), the ankle was placed at 0° of plantarflexion and the ultrasound probe was placed longitudinally, with the middle part of the probe located at 4 cm above the AT’s insertion. A probe holder was used to avoid applying pressure over the tendon. A trial SWE measurement was done to check for possible voids in the stiffness estimation and if voids were detected, the probe was removed, ultrasound gel was added, and the probe was placed again. Elastography images were acquired for 12 seconds (twice). Due to the equipment properties, as a SWE image was obtained every 2.4 seconds; thus, to obtain 4 SWE images, the elastography measurements lasted 12 seconds.

### HD-sEMG and torque recordings

Participants’ skin was shaved, gently abraded (Nuprep, Skin Prep Gel, Weaver and Company, Aurora, Colorado), cleaned with water and dried with tissue paper. Three two-dimensional (2D) adhesive grids (OT Bioelettronica, Italy) of 13 x 5 equally spaced electrodes (each of 1 mm diameter, with an inter-electrode distance of 8 mm) were used to record the HD-sEMG signals from the MG, LG, and SO muscles (position of each grid was described above). The HD-sEMG grid was prepared by attaching a double-sided adhesive foam to the grid surface (SPES Medica, Genova, Italy) and by filling the grid cavities with conductive paste which provided adequate electrode-skin contact (AC-CREAM, SPES Medica, Genova, Italy).

HD-sEMG signals were converted by a 16-bit analogue-digital converter (Quattrocento-OT Bioelecttronica, Torino, Italy). Signals were amplified by a factor of 150, sampled at 2048 Hz, and filtered with a band-pass filter (bandwidth: 10-500 Hz, first order, −3 dB) (Arvanitidis *et al*., 2019). HD-sEMG signals were acquired in monopolar mode with ground electrodes (WhiteSensor WS, Ambu A/S, Ballerup, Denmark) located in the head of the fibula and with a wet strap in the thigh of the assessed leg. Electrode grids and ground electrodes were connected to an bioelectrical amplifier (Quattrocento-OT-Bioelecttronica, Torino, Italy) and the torque exerted by the participants was obtained with a Biodex System 3 dynamometer (Biodex Medical System), which was synchronized with the HD-sEMG signals through the auxiliary input of the EMG amplifier (Arvanitidis *et al*., 2019).

### Image analysis

#### Analysis of Ultrasound images

After obtaining the ultrasound images, a reference line of 1 cm was drawn using the ultrasound device tools. Then, using the software ImageJ (http://imagej.nih.gov/ij), the reference line was converted into pixels, and set as a scale. Next, the width of the ultrasound image was determined, and the middle point marked on the image. To obtain the AT thickness, the distance between the superficial and deep part of the paratenon was measured at 2, 4, and 6 cm from the AT’s insertion. The measurements of three ultrasound images were averaged for each position. Conversely, ultrasound device tools were used to determine the CSA. Briefly, a discontinuous line was drawn following the internal part of the paratenon as reference and the CSA was measured at 2, 4, and 6 cm of its insertion. Similarly, the measurements of three ultrasound images were averaged for each position.

For the stiffness measurements, we obtained approximately 4 SWE color maps (height x width, 2.5 cm x 1 cm) selected using the elastography tools. Then, using the ultrasound device tools, a line was drawn in the middle of the ultrasound image (this line indicates the 4 cm distance from the AT). A region of interest (ROI) of 3 mm diameter (Siu *et al*., 2016) was aligned with the reference line and located in the middle of the tendon, and the stiffness (kPa) was determined. Lastly, mean stiffness was calculated over the ROIs of the 4 consecutive images (Coombes *et al*., 2018).

### HD-sEMG signal analysis

#### Motor unit analysis

The HD-sEMG signals recorded during the two isometric plantarflexion contractions at each target torque (10, 40, and 70% MVC) were decomposed into motor unit spike trains with an algorithm based on blind source separation, which provides automatic identification of multiple single motor units (Martinez-Valdes *et al*., 2022). Single motor units were assessed for decomposition accuracy with a validated metric (Silhouette, SIL), which was set to ≥ 0.90 (Negro *et al*., 2016). SIL is a normalized measure of the relative height of the peaks of the decomposed spike trains with respect to the baseline noise (Martinez-Valdes *et al*., 2022).

The HD-sEMG signals were decomposed throughout the duration of the submaximal contractions, and the discharge times of the identified motor units were converted into binary spike trains (Martinez-Valdes *et al*., 2018). Recruitment threshold was defined as the torque (%MVC) at the time when motor units began firing action potentials and de-recruitment threshold was defined as the torque (%MVC) at the time when motor units ceased firing action potentials (Martinez-Valdes *et al*., 2020). DR and COVisi were calculated during the steady phase of the torque signal (10 s duration). Missing pulses producing non-physiological firing rates i.e., inter-spike intervals > 250 ms, were manually and iteratively deleted and the pulse train was re-estimated. Furthermore, if the algorithm assigned two or more pulses for a single firing, we removed the additional firing, and the final pulse trains were re-calculated (Martinez-Valdes *et al*., 2022). Motor unit data was recorded, analyzed, and reported according to the consensus for experimental design in electromyography: single motor unit matrix (Martinez-Valdes *et al*., 2023).

#### Neuromechanical delay and cross-correlation coefficient

Motor unit rate coding and force generation interactions were determined using cross-correlation to assess delays and similarities between fluctuations in cumulative motor unit firing of each individual muscle (MG, LG, and SO) and torque. Motor unit discharge times obtained were summed to generate a cumulative spike train (CST) that represents the cumulative activity of multiple motor units (Martinez-Valdes *et al*., 2022). CST and torque parameters were low pass filtered (4^th^ order zero-phase Butterworth, 2 Hz) and then high pass filtered (4^th^ order zero-phase Butterworth, 0.75 Hz) as presented previously (Martinez-Valdes *et al*., 2021). Then, CST and torque were cross-correlated to determine the delay in their fluctuations (calculated from the lags obtained from the cross-correlation function) and the similarities in their fluctuations (cross-correlation coefficient) (Martinez-Valdes *et al*., 2022). Delays between the motor unit firing activity and torque were used as a measure of the NMD. Finally, cross-correlation coefficient between signals was computed in 5-s segments with 50% overlap (Martinez-Valdes *et al*., 2022). The average NMD and cross-correlation coefficient calculated from these segments was reported. Furthermore, the NMD and cross-correlation coefficient of the triceps surae muscle (MG, LG, and SO combined) was also calculated.

#### Torque signal analysis

Torque signals were low pass filtered at 15 Hz and then used to quantify torque steadiness (coefficient of variation of torque (COV Torque), SD torque/mean torque * 100) from the steady phase of the contractions (Martinez-Valdes *et al*., 2020). The torque exerted by each participant was plotted using a custom-made MATLAB script allowing us to identify and select the steady phase of the contraction needed for the analysis (Arvanitidis *et al*., 2022).

### Intermuscular coherence and functional muscle network analysis

Intermuscular coherence analysis was performed to identify the presence of a common descending drive shared by motor unit pools of the MG, LG, and SO muscles. Intermuscular coherence is a well-established measure of association of motor unit firing activity between muscles (Dobie & Wilson, 1989; Simpson *et al*., 2000; Miranda de Sá & Felix, 2002). Its calculation generates a connectivity strength value for each pair of EMG signals (in our case motor unit CSTs) within a specified frequency band, resulting in a weighted adjacency matrix for each subject. Significant coherence at low frequencies (<5 Hz) is associated with force production; in the alpha band (5-15 Hz), with involuntary force oscillations (physiological tremor) and afferent feedback loops; in the beta band (15-30 Hz), with corticospinal projections; and in the gamma band (30-60 Hz), with signals relevant to efferent motor commands in the motor cortex (Brown *et al*., 1998). Hence, these four frequency bands were considered in the calculation of the connectivity matrices. Furthermore, to isolate significant connections and exclude less relevant ones, a threshold was applied as has been done previously (Halliday *et al*., 1995). To ensure consistency across subjects, a proportional threshold of 25% was used to maintain the same connection density (Fornito *et al*., 2016).

After obtaining the average coherence matrix for each group, the muscular connectivity was visually represented using graph theory. In this representation, each muscle was depicted as a node within the network, while the functional connections, revealed by intermuscular coherence, were illustrated as edges between these nodes. Graph theory, widely utilized in EEG studies, allows for the analysis of networks with numerous connections and nodes, enabling the application of various metrics for comparison (Fornito *et al*., 2016).

Due to the limited number of muscles accessible for data collection in this study (N=3), the resulting networks have fewer connections and nodes. Consequently, the analysis was centered on the assessment of the strength metric. The strength metric represents the cumulative weights of links connected to a node, indicating the overall strength of connections between muscles. This metric reflects the muscle’s influence or contribution to collective muscle activity, with higher strength values indicating a more prominent role in the neural control of movement (Mheich *et al*., 2020). All network measures were computed using the Brain Connectivity Toolbox, a specialized tool for analyzing functional connectivity and graph-based interactions between signals (Rubinov & Sporns, 2010).

### Statistical analysis

Descriptive statistics were used to report the data which are presented as mean ± SD, unless otherwise specified. The Shapiro-Wilk test was used to determine the normality of the data, and the Levene test was used to evaluate the assumption of homogeneity of variance. As these assumptions were met, parametric statistical tests were deemed appropriate. The level of significance for all statistical procedures was set at P<0.05 and 95% confidence intervals (CI) of differences were reported. Participant’ characteristics, questionnaires scores, morpho-mechanical properties of the AT and connectivity strength were compared between groups using independent T-test for continuous variables. IBM SPSS Statistics software, V. 29.0 (Armonk, New York, USA) was used for these comparisons.

Statistical analyses for the HD-sEMG motor unit data were done using R (version 4.2.1, R Development Core Team, 2023). A generalized linear mixed-effects model (GLMM) analysis was performed with the R package lme4 (version 1.1.31). The following model was used *“variable of interest ∼ group * muscle * torque (1| subject)”.* This model is read as a “variable of interest predicted by each of the factors”. The variable of interest (on the left) is the dependent variable and the ones on the right (except the subject) are the independent (explanatory) variables, or “fixed effects”, group (Control, NIAT), muscle (MG, LG, SO), and torque (10, 40 and 70% MVC). The 1|subject in the model is the “random effect”, which characterizes the variation that is due to individual differences. The normality of residuals was assessed using the Shapiro-Wilk test after running the model. Residual outliers were removed using Cook’s distance method (considering as threshold, 4 times the SD) when the normality assumption was not met. Post-hoc pairwise comparisons were conducted using Sidak correction and least-squares contrasts, as implemented in the R package emmeans (version 1.8.8). The post-hoc tests were used based on the interactions identified with the GLMM. Post-hoc results were reported as p-value and 95% confidence intervals (CI).

Torque steadiness was compared between groups at each torque level with a linear mixed model analysis with factors group (Control, NIAT) and torque (10, 40, 70% MVC) as fixed effects, and participants as random effect using GraphPad Prism software V.8.0.2 (San Diego, California, USA).

Relationships between pain level and questionnaire scores and neuromuscular and morpho-mechanical parameters were analyzed through correlation analysis using IBM SPSS Statistics software, V. 29.0. Pearson correlation coefficients and p-values for each significant correlation were reported.

## RESULTS

### Participants characteristics

All 51 participants (26 NIAT, 25 asymptomatic controls) completed the study. Participants anthropometrics, peak plantar flexion torque and information regarding their symptoms are reported in Table 1. No statistical differences were observed between the groups.

**Table 1.** Participants characteristics.

|  | NIAT group<br>(N=26) | Control group<br>(N=25) | p-value | 95% CI |
| --- | --- | --- | --- | --- |
| Age (years) | 29.0 ± 8.5 | 28.6 ± 3.9 | 0.815 | [-4.17 to 3.30] |
| Sex (males/females) | 14/12 | 17/8 |  |  |
| Height (cm) | 173.0 ± 9.2 | 171.1 ± 9.2 | 0.463 | [-7.08 to 3.27] |
| Weight (kg) | 75.1 ± 17.2 | 74.0 ± 11.6 | 0.793 | [-9.37 to 7.20] |
| Peak torque (Nm) | 85.7 ± 24.7 | 91.6 ± 30.1 | 0.448 | [-9.60 to 21.38] |
| Laterality (Right/Left) | 24/2 | 19/6 |  |  |
| Assessed leg (Right/Left) | 16/10 | 17/8 |  |  |
| Bilateral symptoms | 13/26 | N/A |  |  |
| Symptoms duration<br>(months) | 42.10 ± 38.45 | N/A |  |  |
| NRS | 3.12 ± 1.24 | N/A |  |  |

Values are presented as mean ± SD (Standard deviation). 95% Confidence intervals are reported. NIAT, non-insertional Achilles tendinopathy; NRS, Numerical Rating Scale; N/A, non-applicable. \* Indicates significant statistical difference, $p < 0.05$ .

### Questionnaires

Questionnaire scores between groups are reported in Table 2. Individuals in the NIAT group presented lower scores in the VISA-A (p<0.001), IPAQ-SF (p=0.019), FAAM (Subscale Activities of Daily Living) (p<0.001), and FAAM (Subscale Sports) (p<0.001) compared to the Control group. Additionally, individuals in the NIAT group showed higher TSK scores compared to the Control group (p=0.021). No differences were observed in the PCS scores between groups.

**Table 2.** Questionnaires score.

|  | NIAT group<br>(N=26) | Control group<br>(N=25) | p-value | 95% CI |
| --- | --- | --- | --- | --- |
| VISA-A | 65.73 ± 16.16 | 100 ± 0 | <0.001* | [23.99 to 37.35] |
| IPAQ-SF | 128.46 ± 58.36 | 171.60 ± 68.19 | 0.019* | [7.47 to 78.81] |
| FAAM (Subscale Activities<br>of Daily Living) | 85.42 ± 11.15 | 99.44 ± 1.83 | <0.001* | [9.48 to 18.56] |
| FAAM (Subscale Sports) | 68.15 ± 17.04 | 98.68 ± 3.68 | <0.001* | [23.52 to 37.53] |
| PCS | 12.15 ± 10.65 | 8.72 ± 8.25 | 0.205 | [-8.81 to 1.94] |
| TSK | 36.08 ± 7.64 | 30.80 ± 8.21 | 0.021* | [-9.74 to -0.82] |

Values are presented as mean ± SD (Standard deviation). 95% Confidence intervals are reported. NIAT, non-insertional Achilles tendinopathy; VISA-A, Victorian Institute of Sport Assessment – Achilles Questionnaire; IPAQ-SF, International Physical Activity Questionnaire short form; FAAM, Foot and Ankle Ability Measure, PCS, Pain Catastrophizing Scale; TSK, Tampa Scale for Kinesiophobia. \* Indicates significant statistical difference, $p < 0.05$ .

### Morpho-mechanical properties of the AT

Morpho-mechanical properties of the AT between groups are reported in Table 3. Individuals in the NIAT group presented higher thickness at 6 cm (p=0.034), and CSA at 4 cm (p=0.039) and 6 cm (p<0.001) of the AT compared to Control group. Additionally, individuals in the NIAT group showed lower stiffness of the AT compared to the Control group (p=0.003). No differences were observed in length, thickness at 2 and 4 cm and CSA at 2 cm of the AT between groups.

**Table 3.** Morpho-mechanical properties of the Achilles tendon.

|  | NIAT group<br>(N=26) | Control group<br>(N=25) | p-value | 95% CI |
| --- | --- | --- | --- | --- |
| Length (cm) | 21.23 ± 3.07 | 19.89 ± 2.57 | 0.096 | [-2.94 to 0.25] |
| Thickness 2 cm (cm) | 0.35 ± 0.06 | 0.37 ± 0.04 | 0.400 | [-0.02 to 0.04] |
| Thickness 4 cm (cm) | 0.45 ± 0.10 | 0.41 ± 0.05 | 0.074 | [-0.08 to 0.004] |
| Thickness 6 cm (cm) | 0.47 ± 0.12 | 0.40 ± 0.05 | 0.008* | [-0.12 to -0.02] |
| CSA 2 cm (cm<sup>2</sup>) | 0.46 ± 0.08 | 0.44 ± 0.09 | 0.315 | [-0.07 to -0.02] |
| CSA 4 cm (cm<sup>2</sup>) | 0.44 ± 0.09 | 0.40 ± 0.06 | 0.039* | [-0.09 to -0.002] |
| CSA 6 cm (cm<sup>2</sup>) | 0.48 ± 0.08 | 0.38 ± 0.08 | <0.001* | [-0.14 to -0.05] |
| Stiffness (kPa) | 68.16 ± 7.80 | 75.95 ± 9.98 | 0.003* | [2.76 to 12.82] |

Values are presented as mean ± SD (Standard deviation). 95% Confidence intervals are reported. NIAT, non-insertional Achilles tendinopathy; CSA, cross-sectional area \* Indicates significant
statistical difference, $p < 0.05$ .

### Motor unit decomposition

A total of 5,681 motor units were identified in the triceps surae muscle during submaximal contractions in both the NIAT and control groups. At 10% MVC, the average number of motor units identified in the NIAT group were 14.3 ± 9.2 for MG, 6.5 ± 5.1 for LG, and 15.6 ± 8.4 for SO; in the control group, the averages were 18.9 ± 18.6 for MG, 8 ± 10.9 for LG, and 12.6 ± 8.8 for SO. At 40% MVC, the NIAT group had averages of 25.3 ± 19.5 for MG, 11.4 ± 9.6 for LG, and 16.5 ± 8.0 for SO, while the control group had 25.5 ± 23.4 for MG, 11.1 ± 11.3 for LG, and 12.3 ± 7.5 for SO. At 70% MVC, the NIAT group had averages of 18.5 ± 15.7 for MG, 9.3 ± 11.1 for LG, and 12.1 ± 8.1 for SO, compared to the control group’s 10.4 ± 12.6 for MG, 3.9 ± 2.5 for LG, and 11.9 ± 8.0 for SO.

### Recruitment and de-recruitment threshold

Recruitment thresholds obtained from motor units from MG, LG, and SO muscles at 10, 40, and 70% MVC are presented in **Figure 3A**. Overall, no differences in recruitment threshold were observed between groups across torque levels (Group effect: F=0.56, P=0.46). Within-group muscle differences at each torque level can be seen in **Figure 3A**. De-recruitment thresholds calculated from motor units from MG, LG, and SO muscles at 10, 40, and 70% MVC are presented **in Figure 3B**. In general, de-recruitment threshold increased differently across torque levels between muscles and groups (interaction effect: F=7.24, P<0.0001), with between groups differences showing decreased de-recruitment threshold (i.e., delayed de-recruitment) in the LG in the NIAT group compared to control group at 70% MVC (P=0.039, 95% CI= 0.11 to 10.41). Within-group muscle differences at each torque level can be seen in **Figure 3B**.

**Figure 3.**
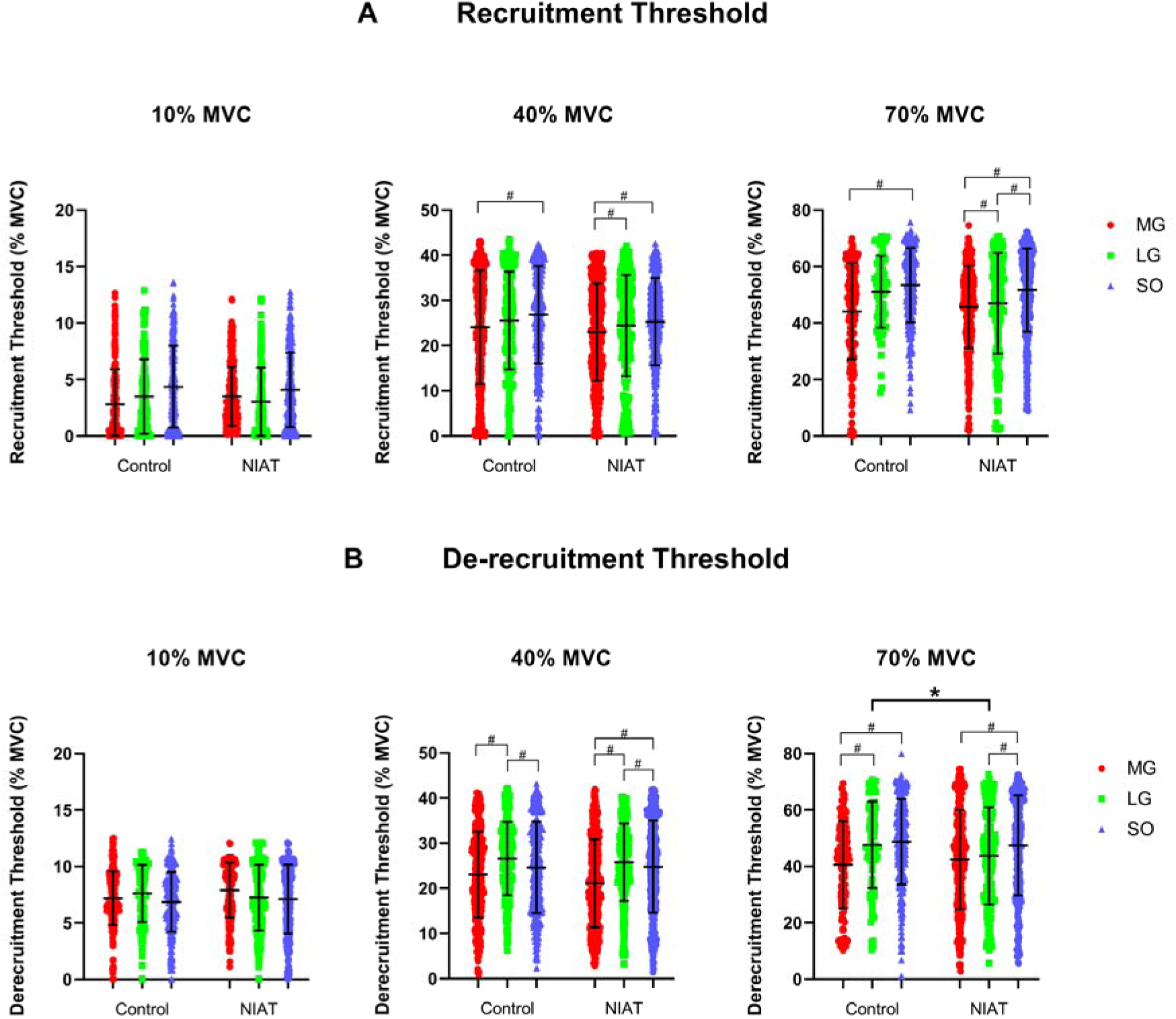
A) Recruitment threshold from medial gastrocnemius (MG: red bar), lateral gastrocnemius (LG: green bar) and soleus (SO: blue bar) muscles at 10, 40, and 70% maximum voluntary contraction (MVC) between control and non-insertional tendinopathy (NIAT) groups. B) De-recruitment threshold from MG, LG, and SO muscles at 10, 40, and 70% MVC between control and non-insertional tendinopathy (NIAT) groups. A linear mixed model was used for statistical comparisons. * Statistical differences between muscles and groups, p<0.05. # Statistical differences between muscles within groups, p<0.05.

### Discharge Rate

DR obtained from motor units from MG, LG, and SO muscles at 10, 40, and 70% MVC are presented in **Figure 4**. Overall, DR increased differently across torque levels between muscles and groups (interaction effect: F=7.52, P<0.0001), with between-group differences showing significantly increased DR in the LG in the NIAT group compared to control group at 70% MVC (P=0.002, 95% CI= −3.27 to −0.34). Within-group muscle differences at each torque level can be seen in **Figure 4**.

**Figure 4.**
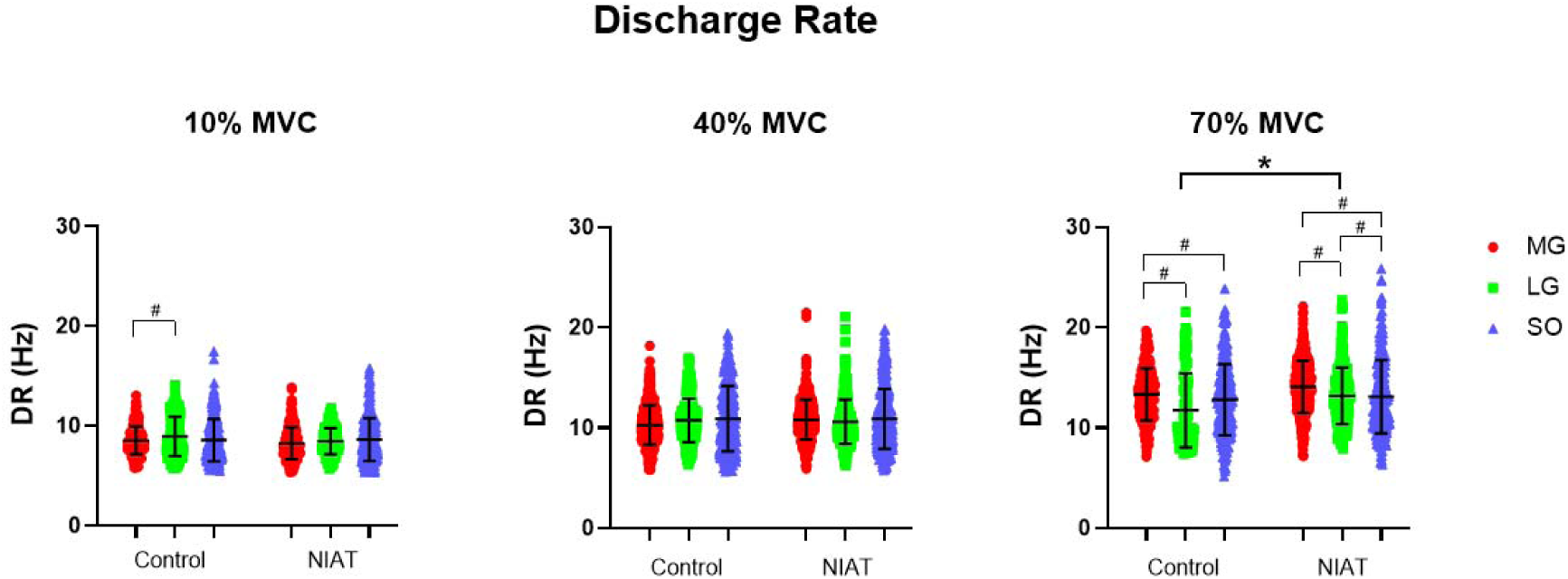
Motor unit discharge rate (DR) from medial gastrocnemius (MG: red dot), lateral gastrocnemius (LG: green square) and soleus (SO: blue triangle) muscles at 10, 40, and 70% maximum voluntary contraction (MVC) between control and non-insertional tendinopathy (NIAT) groups. A linear mixed model was used for statistical comparisons. * Statistical differences between muscles and groups, p<0.05. # Statistical differences between muscles within groups, p<0.05.

### Discharge Rate Variability

COVisi calculated from motor units from MG, LG, and SO muscles at 10, 40, and 70% MVC are presented in **Figure 5**. Overall, COVisi increased differently between groups and muscles (interaction effect: F=4.41, P=0.012); however, no differences between NIAT and control groups were observed. Within-group muscle differences at each torque level can be seen in **Figure 5**.

**Figure 5.**
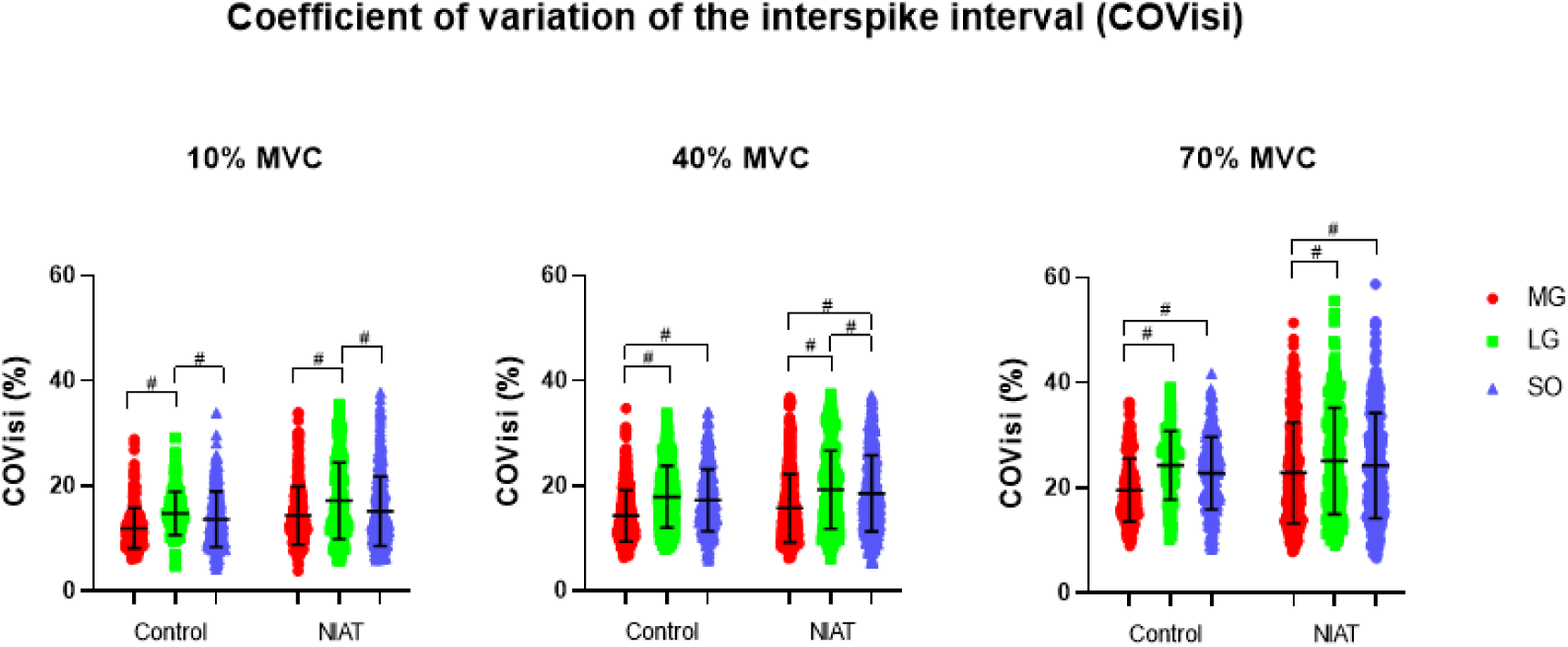
Motor unit coefficient of variation of the interspike interval (COVisi) from medial gastrocnemius (MG: red dot), lateral gastrocnemius (LG: green square) and soleus (SO: blue triangle) muscles at 10, 40, and 70% maximum voluntary contraction (MVC) between control and non-insertional tendinopathy (NIAT) groups. A linear mixed model was used for statistical comparisons. # Statistical differences between muscles within groups, p<0.05.

### Neuromechanical delay and cross-correlation coefficient

NMD calculated from motor units from MG, LG, and SO muscles at 10, 40, and 70% MVC are presented in **Figure 6A**. Overall, NMD decreased distinctly across torque levels between muscles and groups (interaction effect: F=11.74, P<0.0001); however, no differences between NIAT and control groups were observed. Within-group muscle differences at each torque level can be seen in **Figure 6A**. CST and torque cross-correlation coefficient calculated from motor units from MG, LG, and SO muscles at 10, 40, and 70% MVC are presented in **Figure 6B**. In general, cross-correlation coefficients varied across torque levels between muscles and groups (interaction effect: F=84.27, P<0.0001), with between groups differences showing significantly decreased cross-correlation coefficient in the LG in the NIAT group compared to control group at 10% MVC (P<0.0001, 95% CI= 0.07 to 0.27) and increased cross-correlation coefficient in the LG in the NIAT group compared to control group at 70% MVC (P=0.013, 95% CI= −0.21 to −0.01). Within-group muscle differences at each torque level can be seen in **Figure 6B**.

**Figure 6.**
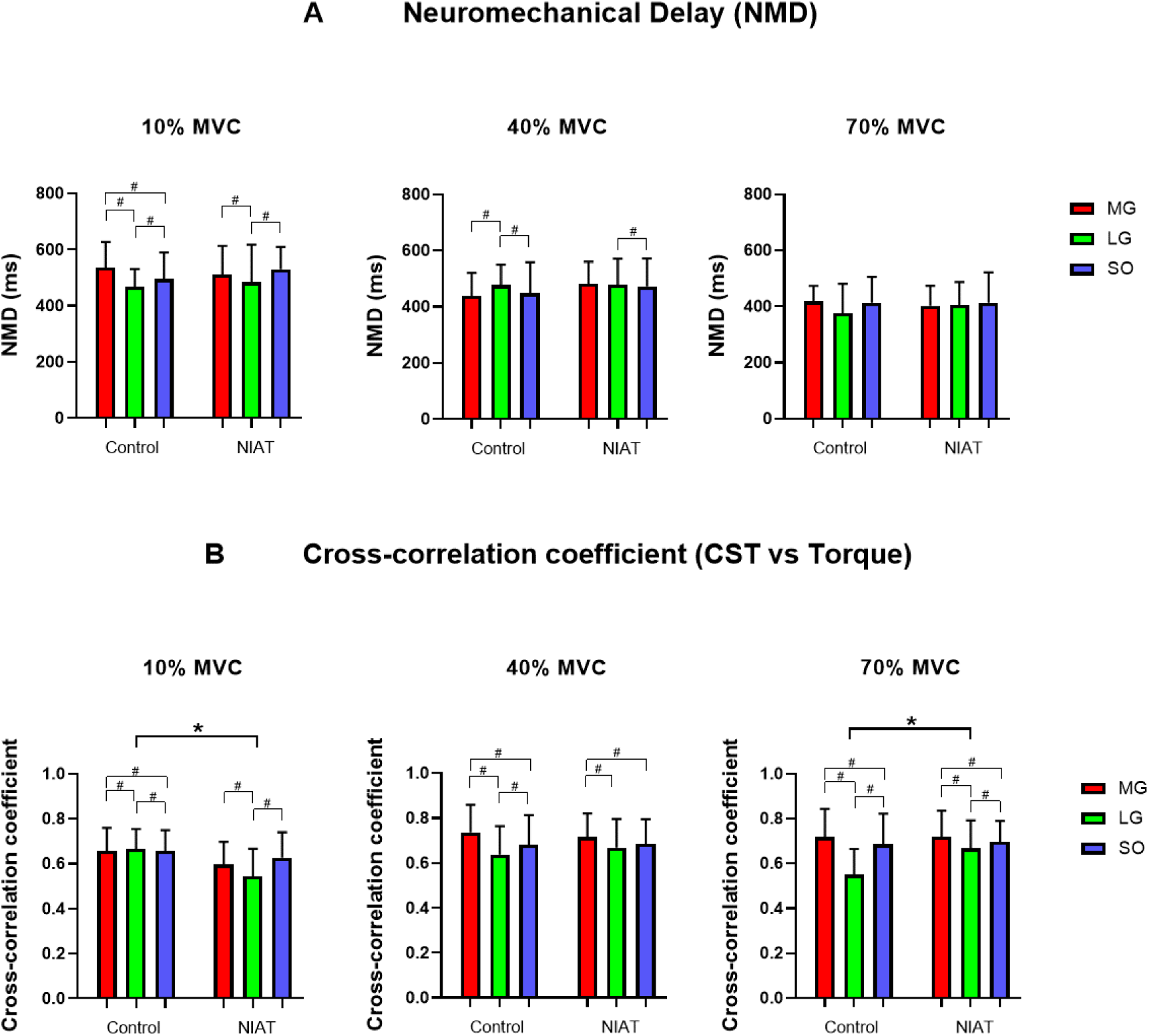
A) Neuromechanical delay (MND) from medial gastrocnemius (MG: red bar), lateral gastrocnemius (LG: green bar) and soleus (SO: blue bar) muscles at 10, 40, and 70% maximum voluntary contraction (MVC) between control and non-insertional Achilles tendinopathy (NIAT) groups. B) Cross-correlation coefficients between cumulative spike train (CST) vs torque from MG, LG, and SO muscles at 10, 40, and 70% MVC between control and NIAT groups. A linear mixed model was used for statistical comparisons. * Statistical differences between muscles and groups, p<0.05. # Statistical differences between muscles within groups, p<0.05.

### Torque steadiness

Overall, torque steadiness varied across torque levels (Torque effect: F=29.94 P<0.0001, MD=0.28, 95% CI= −0.04 to 0.60); however, no differences between groups were observed (Group effect: P=0.09).

### Connectivity strength and functional muscle networks

The connectivity strength and functional muscle networks derived from intermuscular coherence analysis for the 0-5 Hz frequency band, calculated from the CSTs of the MG, LG, and SO muscles at 10% MVC, are shown in **Figure 7A**. Individuals in the control group presented higher connectivity strength compared to the NIAT group (p=0.03). Functional muscle networks (MG:M1, LG:M2, and SO:M3) showed decreased connectivity between MG and LG muscles in the NIAT group compared to the control group. Connectivity strength for the frequency band 5-15 Hz obtained from the CSTs of the MG, LG, and SO muscles at 10% MVC can be seen in **Figure 7B**. Individuals in the control group showed higher connectivity strength in this bandwidth compared to the NIAT group (p<0.001). Functional muscle networks showed decreased connectivity between MG and LG muscles in the NIAT group compared to the control group. No differences in connectivity strength for the frequency bands 15-30 Hz and 30-60 Hz were observed between control and NIAT groups at 10% MVC. Additionally, no differences in connectivity strength for the frequency bands 0-5 Hz, 5-15 Hz, 15-30 Hz, and 30-60 Hz were observed between control and NIAT groups at 40 and 70% MVC (p>0.09 in all cases).

**Figure 7.**
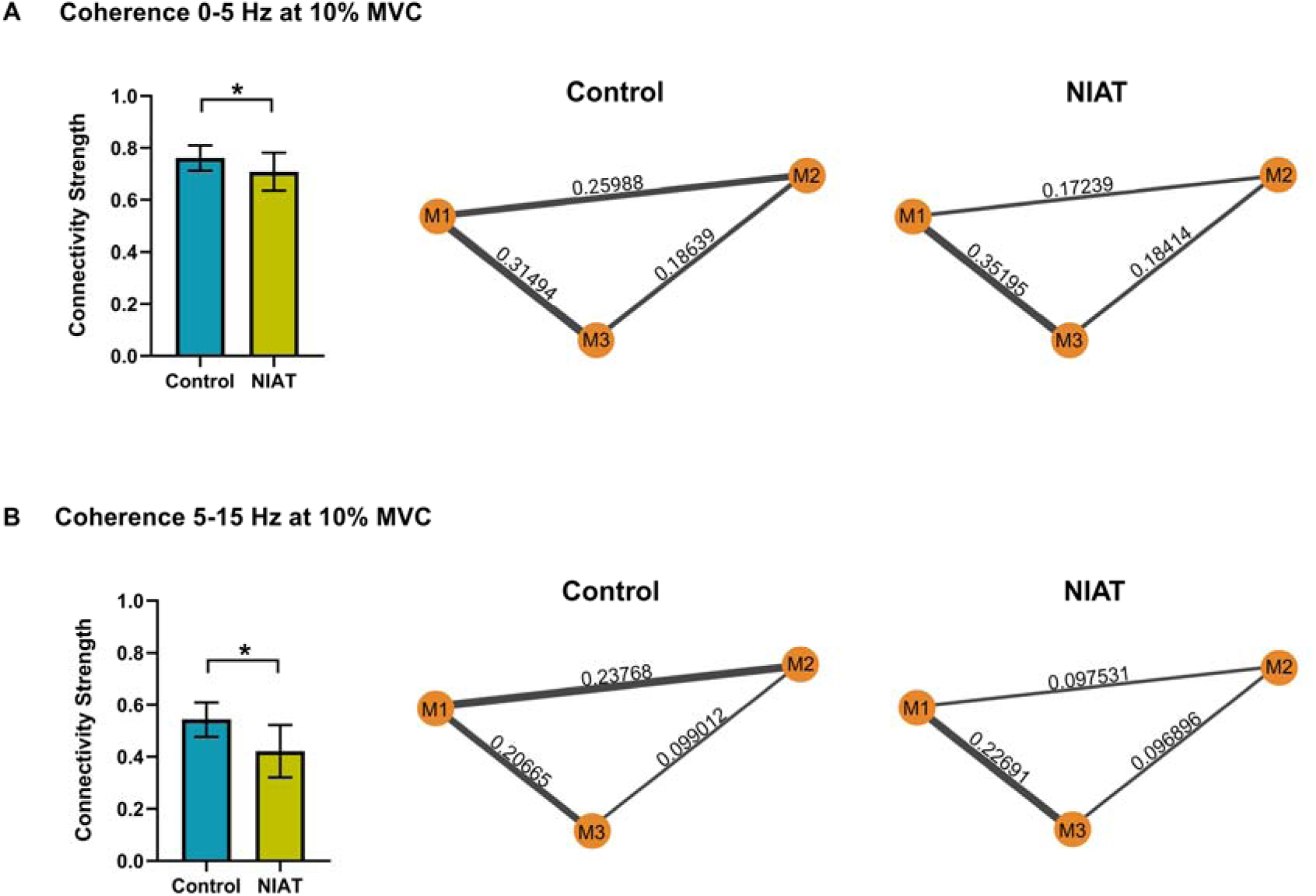
A) Left, intermuscular coherence for the frequency band 0-5 Hz at 10% maximum voluntary contraction (MVC) between control and non-insertional Achilles tendinopathy (NIAT) groups. Right, functional muscle networks for the medial gastrocnemius (MG:M1), lateral gastrocnemius (LG:M2), and soleus (SO:M3) muscles between control and NIAT groups. B) Left, intermuscular coherence for the frequency band 5-15 Hz at 10% MVC between control and NIAT groups. Right, functional muscle networks for the MG, LG, and SO muscles between control and NIAT groups. An unpaired t-test was used for statistical comparisons. * Statistical differences between groups, p<0.05.

### Relationships between pain level and questionnaires and neuromuscular and morpho-mechanical parameters in individuals with NIAT

A correlation analysis was conducted to assess the relationship between pain level and questionnaires and neuromuscular and morpho-mechanical parameters. The pain level was significantly negatively correlated with LG’s de-recruitment threshold at 70% MVC (r= −0.52, P<0.05). VISA-A was significantly negatively correlated with AT thickness 6 cm (r= −0.46, P<0.01). Moreover, FAAM (Subscale Activities of Daily Living) was significantly negatively correlated with AT thickness 6 cm (r= −0.64, P<0.001) and CSA 6 cm (r= −0.42, P<0.05). Finally, PCS was significantly positively correlated with AT thickness 6 cm (r=0.49, P<0.05). Additionally, there were no significant correlations between the IPAQ-SF, FAAM (Sports Subscale), and TSK questionnaires with motor unit properties or the tendon’s morpho-mechanical parameters.

## DISCUSSION

This study demonstrates that individuals with NIAT exhibit load- and muscle-dependent alterations in motor unit discharge rate, motor unit firing-torque relationships, and intermuscular coherence of the triceps surae during isometric plantarflexion contractions. Specifically, our findings show that, at higher force levels, individuals with NIAT display an increased discharge rate and a reduced de-recruitment threshold in the LG compared to controls. Additionally, the results reveal that the cross-correlation coefficient between motor unit activity and torque production (CST vs. Torque) in the LG is lower at low forces and higher at high forces in individuals with NIAT, suggesting an altered contribution of the LG to the overall plantarflexion torque. Moreover, our analysis indicates a reduced connectivity strength within the triceps surae for low-frequency bands (0-5 and 5-15 Hz) at low forces in individuals with NIAT. Notably, the functional muscle network analysis highlights a decreased connectivity between the MG and LG muscles, possibly reflecting an imbalance in the synergistic activity of the triceps surae in individuals with NIAT.

### Differential changes in motor unit firing properties for MG, LG, and SO muscles in individuals with NIAT

There is evidence that some muscles are predominantly controlled by a common neural drive (Bremner *et al*., 1991; Gibbs *et al*., 1995; Laine *et al*., 2015; Kerkman *et al*., 2018), with the strength of this shared drive being greater among muscles closely related anatomically and functionally (Gibbs *et al*., 1995; Kerkman *et al*., 2018). However, recent evidence indicates that the common neural drive between the MG, LG, and SO muscles is reduced during isometric plantarflexion contractions and heel-raise tasks (Hug *et al*., 2021). Our findings are in accordance with those results since we observed muscle differences in recruitment and de-recruitment thresholds, DR, COVisi, NMD and cross-correlation coefficients at different torque levels, providing further evidence of the limited shared neural drive to the triceps surae muscles in both healthy individuals and those with NIAT. However, we also observed specific motor unit firing changes for the NIAT group in the LG muscle. The NIAT group showed increased DR and decreased de-recruitment thresholds (as illustrated in Fig. 4 and Fig. 3 B, respectively) at 70% MVC, revealing a load-dependent adaptation in motor unit firing properties for this muscle, specifically.

To our knowledge, only one study has investigated the motor unit firing properties of the triceps surae in individuals with NIAT. Consistent with our findings, Fernandez et al. (2022) reported no significant differences in DR of the MG, LG, and SO muscles between NIAT and control groups during isometric plantarflexion contractions at 10% and 20% of the MVC. However, they observed that in the NIAT group, the DR of the LG remained unchanged as the target torque increased from 10% to 20% MVC. Contrary to this, our study found no such blunted firing rate response in the LG; instead, we observed a more pronounced increase in LG’s DR at higher force levels. Additionally, we noted a reduction in the de-recruitment threshold of the LG at 70% MVC. Several factors may explain these observations. First, NIAT is known to alter the mechanical properties of the Achilles tendon, potentially requiring greater motor unit firing to maintain the necessary torque as a compensatory mechanism for the reduced tendon stiffness. This hypothesis is supported by De la Fuente et al. (De la Fuente *et al*., 2023), who observed increased activation of the MG muscle at high force levels (90% MVC) in individuals post-Achilles tendon rupture repair—an injury that increases tendon compliance. Indeed, changes in tendon mechanical properties can alter force transmission and movement generation. However, it is crucial to consider why only the LG exhibits this increased DR at higher forces. NIAT is also associated with increased tendon pain, which may alter the plantarflexion force vector, leading to greater recruitment of the LG muscle at high forces. This idea is supported by the findings of Hug et al. (Hug *et al*., 2021), who demonstrated that the mean discharge rate of the LG can be modulated by changes in foot position during plantarflexion. Therefore, it is possible that NIAT-induced pain-related motor adaptations result in increased neural drive to the LG at higher forces. This pain-related adaptation is further evidenced by the observed reduction in de-recruitment threshold at high forces. Martinez-Valdes et al. (Martinez-Valdes *et al*., 2020) previously showed that experimental muscle pain reduces the recruitment and de-recruitment thresholds of higher-threshold motor units, increasing the duration of their activity during contraction. These higher-threshold motor units are associated with greater force production but have reduced resistance to muscle fatigue. Consequently, the reduced de-recruitment threshold observed in our study might suggest that the increased activation of the LG during the task could lead to inefficient recruitment within the triceps surae group. Typically, the LG exhibits a higher recruitment threshold compared to the MG and SO during plantarflexion (Hug *et al*., 2021). Thus, prolonged overactivation of the LG could potentially increase fatigue in the triceps surae due to altered motor strategies in response to pain, which may exacerbate symptoms over time. Future studies should investigate potential associations between pain and fatigue in individuals with NIAT to further understand these adaptations.

### NMD and CST-torque cross-correlation coefficients

The conversion of neural signals to force output has a latency caused by the dynamic responsiveness of motor neurons and the time required to stretch the series elastic components of the muscle-tendon unit after the activation of the muscle fibers (Del Vecchio *et al*., 2018). In this context, the NMD represents the time difference between the neural drive and the generated torque during a voluntary contraction and can be estimated from the time lag of the peak of the cross-correlation between the CST and torque (Del Vecchio *et al*., 2018). Our results showed no differences in NMD of the MG, LG, and SO between the NIAT group and control group at 10%, 40%, and 70% MVC; however, NMD decreased as the target torque increased in both groups. The decrease in the NMD as the target torque increase may be explained by the Henneman’s size principle (Henneman, 1957), since as the target torque increases larger motor units, which innervate fast-twitch muscle fibers, are recruited. Consequently, the recruitment of fast-twitch muscle fibers may contribute to the observed decrease in NMD. Theoretically, the stiffness of the AT affects the time to stretch the elastic components of the muscle-tendon unit during a voluntary contraction. Given our observation of reduced tendon stiffness in individuals with NIAT, we anticipated that this difference might influence NMD results. However, no significant differences between the NIAT and control groups were observed. In contrast, a previous study by Chang and Kulig (Chang & Kulig, 2015) reported an increase in electromechanical delay (quantified as the time difference between the onset of force/torque and muscle activation during electrically induced contraction) in individuals with NIAT. These conflicting findings may be partially explained by differences in how NMD and electromechanical delay are calculated. Electromechanical delay is typically estimated at a single time point when the muscle activates from a passive state (Chang & Kulig, 2015). In contrast, NMD is influenced by active motor unit responses, such as twitch duration and afferent inputs, during contraction and is measured when the tendon is under sustained tension, reflecting a state of greater tendon stiffness. It is possible that delays between motor unit/EMG activity and torque are more pronounced in NIAT during the initial phase of the force-tension curve, particularly at the contraction intensities generated by electrically induced contractions. However, testing this hypothesis would require a simultaneous assessment of motor unit firing properties and tendon stiffness changes across the full force-tension curve of the tendon.

Recent research has established that CST can be used to estimate the effective neural drive received by a group of synergistic muscles in humans (Mazzo *et al*., 2021). Mazzo and colleagues demonstrated that the neural drive to the triceps surae muscle group (via CST analysis) is correlated with fluctuations in isometric plantarflexion torque (Mazzo *et al*., 2022). Therefore, assessing CST-torque relationships for individual heads of the triceps surae can provide insight into their independent contributions to the resultant plantarflexion torque. Our results indicate that the cross-correlation coefficient between CST and torque decreased in the lateral gastrocnemius (LG) in the NIAT group compared to the control group at 10% MVC and increased in the NIAT group at 70% MVC. These findings suggest a reduced contribution of the LG to net torque at low forces in individuals with NIAT, consistent with previous research that indirectly showed a significant reduction in LG’s contribution to plantarflexion torque at low forces in this population. However, the same study also reported a decreased contribution of the LG at intermediate forces, a difference we did not observe. This discrepancy may be partly due to methodological differences, as Crouzier et al. (2020) used LG activation and relative physiological cross-sectional area to indirectly estimate the muscle’s contribution to net plantarflexion torque. Since surface EMG can be influenced by muscle properties, any change in LG morphology induced by NIAT can alter sEMG calculations and therefore affect individual muscle force estimations, particularly at higher forces due to amplitude cancellation (Dideriksen & Farina, 2019). Furthermore, Crouzier et al. did not assess force levels as high as the ones used in the present study. The observed decrease in LG’s CST-torque cross-correlation at low forces and the increase in cross-correlation at higher forces is noteworthy. As previously discussed, these different force levels exert varying degrees of tension on the tendon. The tension produced at 10% MVC likely demands greater muscular control due to reduced tendon stiffness, which may necessitate a more balanced contribution from all triceps surae muscle heads to the resultant torque. Indeed, at this lower force level, despite significant muscle differences, the control group exhibits more consistent CST-torque cross-correlation coefficients compared to the NIAT group. Interestingly, this pattern reverses at higher forces, where the NIAT group demonstrates a greater contribution of LG to the resultant torque compared to the control group. Given that both MG and SO have larger CSAs and contribute more significantly to the resultant plantarflexion torque (Maganaris, 2003) compared to LG, it would be reasonable to expect that individuals in the control group predominantly rely on these muscles to increase plantarflexion torque. However, this pattern was not observed in the NIAT group. As mentioned in the discussion of the DR results, it is plausible that the NIAT group employed different plantarflexion strategies at various force levels in response to pain. Since tendinopathic pain typically intensifies with increased load (Magnusson *et al*., 2010), the NIAT group may have adjusted the plantarflexion torque vector to reduce the engagement of the MG and SO muscles, thereby attempting to minimize the load on a potentially painful tendon region. However, this hypothesis should be validated in future studies that assess both overall plantarflexion torque and plantar pressure distribution.

### Strength of synergistic activity in individuals with NIAT

When the CNS initiates a voluntary movement, it triggers the activation of several muscles through the simultaneous activation and coordination of thousands of motor units (Bizzi & Cheung, 2013). It has been proposed that the CNS simplifies this task by controlling groups of muscles, known as muscle synergies, instead of commanding each muscle independently (Singh *et al*., 2018). Thus, the CNS encodes several muscle synergies, and it combines them according to a specific task, generating the muscle contractions necessary for the desired movement (Alessandro *et al*., 2013). Intermuscular coherence analysis is a well-established method for identifying shared neural drive, as it quantifies the frequency spectrum of correlated activity between motor units from different muscle pairs (Laine *et al*., 2021). For this reason, we employed intermuscular coherence together with network analysis, to assess the strength of the neural connectivity (shared neural drive) between the three triceps surae muscle heads. Our network analysis demonstrated a decrease in the connectivity strength within the 0-5 Hz band (delta band) for the triceps surae in individuals with NIAT at 10% MVC. This decrease in connectivity strength was primarily given by a reduction in intermuscular coherence between MG and LG muscles. Motor unit coherence at frequencies below 5 Hz reflects concurrent fluctuations in motor unit firing rates (common neural drive) between muscles (De Luca *et al*., 1982; Myers *et al*., 2004) and plays a crucial role in controlling muscle force (Farina & Negro, 2015). These findings, coupled with the reductions in LG’s CST-torque cross-correlation estimates, further support the evidence of reduced LG involvement in generating plantarflexion torque at low forces in individuals with NIAT.

Additionally, our findings showed a decrease in the connectivity strength within the frequency band 5-15 Hz (alpha band) for the triceps surae at 10% MVC in individuals with NIAT. Again, observing a decrease of the intermuscular coherence between MG and LG muscles. Since motor unit coherence at frequencies between 5-15 Hz has been associated with involuntary force oscillations (physiological tremor) (Laine *et al*., 2015), it can be hypothesized that these oscillations might be transmitted to the Achilles tendon and subsequently influence torque steadiness. However, our findings did not reveal any differences in torque steadiness between NIAT and control groups across torque levels. Considering that increased alpha-band intermuscular coherence is typically linked to tasks demanding a high level of muscle coordination (Nazarpour *et al*., 2012; de Vries *et al*., 2016; Laine & Valero-Cuevas, 2017; Reyes *et al*., 2017), and given that the task in our study required precise matching of plantarflexion torque to visual feedback, it is plausible to speculate that individuals with NIAT may experience altered coordination between the MG and LG muscles during low-force contractions, potentially due to reduced afferent input. Indeed, literature suggests that coordinated patterns of this frequency band may reflect afferent feedback loops (arising from 1a or 1b fibers), which is supported by observations that cycles of excitation around the monosynaptic stretch reflex can induce oscillations in the 5-15 Hz frequency band within individual muscles (Lippold, 1970; Christakos *et al*., 2006; Erimaki & Christakos, 2008). Additionally, it has been proposed that the oscillations in the 5-15 Hz frequency band may be linked to the activity of the cerebello-thalamo-cortical-circuit, which resonates at these frequencies and synchronizes with motor output via its projections to the cortex and/or pontomedullary reticular formation (Gross *et al*., 2002; Soteropoulos & Baker, 2006; Williams *et al*., 2010).

### Relationships between pain level and questionnaires scores and neuromuscular and morpho-mechanical parameters

Our results showed a negative correlation between pain level and LG de-recruitment threshold at 70% MVC, suggesting that as the pain level increases, individuals with NIAT decrease the de-recruitment threshold of the LG at high forces, allowing them to maintain the LG active for a greater period to accomplish the task. Additionally, we found a negative correlation between VISA-A and AT thickness 6 cm, indicating that as the tendon thickness increases, the severity of the symptoms increases. Nevertheless, previous studies have shown inconsistent relationships between the extent and/or severity of tendon abnormalities and severity of symptoms (Martin *et al*., 2018). Similarly, we observed a negative correlation between FAAM (Subscale Activities of Daily Living) and AT thickness 6 cm and CSA 6 cm, showing that as the tendon thickness and cross-sectional area increase, the physical function of individuals with NIAT decreases, providing additional evidence of the relationship between tendon morphology and physical function. Finally, we found a positive correlation between PCS and AT thickness at 6 cm, revealing that as the tendon thickness increases, pain catastrophizing (rumination, magnification, and helplessness about pain) increases. Since morphological changes to the AT in individuals with NIAT are commonly linked to the duration of symptoms, our results support the idea that chronic NIAT is linked to changes in psychological attitudes about pain. Understanding the interplay between symptoms, functionality, psychological attitudes towards pain and the neuromuscular and morpho-mechanical properties is essential for an enhanced comprehension of the different factors involved in NIAT. Designing intervention strategies that consider the dynamic interplay between these components is imperative to achieve optimal outcomes in the management of this condition.

### Limitations and future developments

There are some methodological aspects of this study which should be considered.

First, tendon stiffness measurements were conducted under resting conditions. Unfortunately, assessing tendon stiffness with SWE during plantarflexion contractions presents significant challenges, mainly due to foot movement within the isokinetic ankle attachment. This movement introduces artifacts in the elastography color map, which compromises the accuracy of stiffness estimations. Additionally, current SWE devices have very low sampling resolution (e.g., up to 2 SWE images per second) and do not reliably estimate tendon stiffness during contractions (Khair *et al*., 2024). Future studies should utilize more advanced SWE devices to simultaneously assess AT stiffness and triceps surae motor unit firing rates during plantarflexion contractions, offering a more comprehensive understanding of the interplay between neural coding and muscle-tendon behavior in this condition. Furthermore, given that recent studies have successfully identified motor units during both shortening and lengthening contractions (Glaser & Holobar, 2019; Oliveira & Negro, 2021), future research should explore triceps surae motor unit firing parameters during dynamic plantarflexion contractions in individuals with NIAT.

## CONCLUSION

This study demonstrates that individuals with NIAT exhibit load- and muscle-dependent changes in triceps surae motor unit discharge rate, motor unit firing-torque relationships, and intermuscular coherence during isometric plantarflexion contractions. Our research found increased DR and decreased de-recruitment thresholds in the LG at high forces, indicating an altered neural drive to the triceps surae in individuals with NIAT. Additionally, the results revealed a decreased cross-correlation coefficient (CST vs. Torque) in the LG at low forces, with an increase at high forces, suggesting a shift in the LG’s contribution to net plantarflexion torque. Furthermore, our findings indicated reduced strength connectivity of the triceps surae in low-frequency bands at low forces, pointing to a more unbalanced synergistic activity within the triceps surae in individuals with NIAT. Collectively, these results support the hypothesis that the LG may play a central role in the pathophysiology of this condition, potentially affecting load transmission to the Achilles tendon.

## ACKNOWLEDGEMENTS

We thank Joeri van Helden for his assistance during data collection. All figures were created using BioRender.

## DATA AVAILABILITY STATEMENT

The data and analysis codes are available from the corresponding author, EM-V, upon reasonable request.

## AUTHOR CONTRIBUTIONS

IC-H, EM-V, and DF conceived and designed research; IC-H and MA performed experiments; IC-H, DJ-G, and EM-V analysed data; IC-H and EM-V interpreted results of experiments; IC-H prepared figures; IC-H and EM-V drafted manuscript; IC-H, EM-V, DF, FN, DJ-G and MA edited and revised manuscript; IC-H, EM-V, DF, FN, DJ-G and MA approved final version of manuscript.

